# Influence of a Dietary Supplement on the Gut Microbiome of Overweight Young Women

**DOI:** 10.1101/2020.02.26.20027805

**Authors:** Peter Joller, Sophie Cabaset, Susanne Maurer

## Abstract

**Background:** The importance of the gut microbiome cannot be overstated. Besides the help in digestion, it influences the immune system, produces immunologic and neurologic communication molecules, hormones and vitamins. The gut microbiome is also involved in controlling the BMI (Body Mass Index).

**Objectives:** We attempted to influence the gut microbiome with a herbal yeast-based dietary supplement in 26 women (BASEC ID 2016-02162; ClinicalTrials.gov NCT03223987, BS2016.1) with a BMI between 30 and 35 and and aged 25 to 35 years within 3 weeks. The clinical trial was conducted in accordance with the Declaration of Helsinki. Our hypothesis was that the supplement modifies the microbiome individually by more than 20%.

**Methods:** With shotgun sequencing in the 2 stool samples of each participant we found 68 bacterial species.

**Results:** The microbiome was altered by more than 20%. Five bacterial species were identical to probiotic species known as good butyrate producers and described as beneficial for the gut. Seventy percent of our study cohort showed an increase in the majority of these beneficial microbes during the study. The ratio of Firmicutes to Bacteroidetes is an important parameter in analyzing overweight persons. Twelve of the participants initially showed a F/B ratio (Firmicutes to Bacteroidetes ratio) above 1.6. After three weeks, five of these women normalized.

**Conclusions:** A baker’s yeast-based food supplement is able to modulate the gut microbiome of overweight young women within 3 weeks.

**Lay Summary:** Twenty-six overweight women 25 to 35 years old took an alimentary supplement consisting of plasmolyzed herbal baker’s yeast during three weeks. The goal was to influence the gut bacteria (microbiome). In the stool samples by molecular techniques we could show that the amount of individual bacteria in our cohort changed by more than 20% within the 3 weeks. Some the bacteria found are described as beneficial in the scientific literature. Over 70% of the participants showed an increase in the majority of these advantageous microbes.

Firmicutes and Bacteroidetes are the two major phyla in our microbiome. The ratio of these phyla is associated with the body weight. The more Bacteroidetes, the lower the ratio and the body weight. During ingestion of the supplement, in 14 participants the ratio became lower.

## Introduction

The gut microbiota is a highly important entity of the human body (Jandhyala et al., 2015), (Marchesi et al., 2016). The human intestine contains the largest community of the roughly 100 trillion (Sender et al., 2016) commensal and symbiotic bacteria (Marietta et al., 2015). It influences the immune system (Thaiss et al., 2016), autoimmunity (Shamriz et al., 2016), allergies (Prince et al., 2015), inflammatory bowel disease (Frank et al., 2011), depression (Liśkiewicz et al., 2019), the wellbeing (Neto & O’Toole, 2016) and the BMI (Chen et al., 2014),(Goodrich et al., 2014). The gut-associated lymphoid tissue (GALT) is our largest immunologic organ with over 70% of the total immune system.(Vighi et al., 2008) Over the *Nervus vagus*, there is a direct bi-directional connection between the gut and the brain (Galland, 2014),(El Aidy et al., 2016). Newer publications find a link between the gut microbiome and obesity (Bertram et al., 2017), (Bischoff, 2017), (Weghuber, 2019), Attention Deficiency Syndrome (Stevens et al., 2019),(Aarts et al., 2017),(Shivani Ghaisas, Joshua Maher, 2012), personalized medicine (Doestzada et al., 2018),(Kinross et al., 2011), Alzheimer’s disease (Hu et al., 2016) and even autism (Mulle et al., 2013), (Kang et al., 2017), (Li et al., 2017). It was found that sex hormones, BMI and dietary fiber contribute to shaping the gut microbiome in humans (Dominianni et al., 2015),(Wu et al., 2011). The gut microbiome is considered as a human organ with its own specific functions and complexity (O’Hara & Shanahan, 2006),(Baquero & Nombela, 2012),(Manson et al., 2008). The microbiome produces hormones, neurotransmitters, immunologic signal molecules, SCFA (Short-Chain Fatty Acids) and mucus. A human or a mammalian life without the gut microbiome is not conceivable. But it would be wrong to attribute only beneficial effects to the gut bacteria (Mills et al., 2019). Depending on the number of bacteria and the composition of the microbiome it can also produce dysbiosis, toxic effects, inflammation, and infectious diseases. The gut bacteria exhibit more than three million genes producing thousands of metabolites, whereas the human genome has only about 23’000 genes (Valdes et al., 2018). In contrast to the microbiomes of children or elderly people, in persons between 20 and 65 years of age, the microbiome is stable (Lozupone et al., 2012), (Faith et al., 2013). This temporal stability, however, can be broken by disease, antibiotics or diet (David et al., 2014). Until now there is a lack of knowledge about a “normal” microbiome. Therefore the Human Microbiome Project (HMP, http://www.hmpdacc.org) was instituted in 2007 (Gevers et al., 2012).

Since it is known that diet can directly influence the composition of the gut microbiota (Conlon & Bird, 2015),(Kau et al., 2011),(Davenport et al., 2014),(Voreades et al., 2014), (Turnbaugh et al., 2009), (Laitinen & Mokkala, 2019), and inspired by publications stating that yeast β-glucan and α-mannan are considered beneficial components of foods (Nakashimada et al., 2011), (Cuskin et al., 2015),(Wegmann et al., 2014) we designed a study to find out whether a baker’s yeast-based dietary supplement is able to alter the composition of the gut microbiome in probands in a clinically relevant dimension within three weeks. Our working hypothesis was that following the course of a three-weeks supplemention regimen a shift of 20% in most of the different bacterial species of the microbiome could be expected. The number of the probands was determined accordingly. We also wanted to know, if a potential shift in the bacterial composition is considered beneficial or not.

## Methods

### Study design

#### Participants

26 healthy women aged 25-35 with a measured body mass index (BMI, kg/m^2^) between 30 and 35 were recruited to a specialized medical center (Adimed - Zentrum für Adipositas-und Stoffwechselmedizin Winterthur, Switzerland). After approval of the competent ethics committee (BASEC ID 2016-02162; ClinicalTrials.gov NCT03223987, BS2016.1) and written informed consent by the participants, the probands were assessed for the inclusion and exclusion criteria. The inclusion criteria were female gender; age (between 25 and 35 years); BMI (between 30 and 35 kg/m2); fluency in German language and smartphone users. The exclusion criteria were the presence of nutrition therapy dependent diseases and other serious diseases requiring continuous drug therapy; women who underwent a diet during the last 6 months, took drugs for weight loss at any time before the study, or were enrolled in any weight loss program; regular consumption of pre-or probiotics. For statistical reasons and in an effort to minimize bias we attempted a homogenous group.

### Study conduct

This study was executed as a 5-weeks single-arm pre/post pilot intervention. Recruited participants attended three visits at Adimed. During their first visit, the terms and conduct of the study were explained to the study participants. The recruited women were then asked to sign an informed consent before undergoing a physical examination. At the end of the consultation, participants were given the required tubes for stool sampling. The second visit took place right before the start of the intervention: the participating women brought back their first stool sample and were given the dietary supplement, which they were asked to take during three weeks. Finally, the third visit took place at the end of the intervention. Participating women brought back their second stool sample and the remaining quantity of their dietary supplements. Compliance was measured by assessing the quantity of dietary supplement remaining after three weeks of intervention.

### Stool Sample collection

Stool samples were collected at home in an OMNIgene*®*·GUT tube (DNA Genotek Inc. 3000 -500 Palladium Drive, Ottawa, ON, Canada))(Ilett et al., 2019).

### Dietary supplement

Strath® liquid is a plasmolyzed herbal yeast preparation with over 60 vital substances (such as vitamins, minerals, enzymes, amino-acids, oligo-elements), sold worldwide for almost 60 years (Bio-Strath® AG, Zurich, Switzerland, https://bio-strath.com/). Ingredients: 83% Plasmolyzed Herbal Yeast (type: Saccharomyces cerevisiae MEYEN), 9% Malt extract, 5% Orange syrup, 3% Honey. Daily intake in the study: 3 × 5ml for three weeks.

All study procedures were approved by the local institutional review board and are registered under ClinicalTrials.gov

### Shotgun Metagenome Sequencing (SMS)

The extracted stool samples were analyzed by shotgun sequencing (CoreBiome, St. Paul, MN, USA).

### DNA Extraction

Samples were extracted with Qiagen DNeasy 96 PowerSoil Pro automated for high throughput on QiaCube (Qiagen, Germantown, MD, USA). DNA Quantification: Samples were quantified with Qiant-iT Picogreen dsDNA Assay (Life Technologies Corporation, Grand Island, NY, USA). All samples passed the internal quality control.

Library Preparation and Sequencing: Libraries were prepared with a procedure adapted from the Nextera Library Prep kit (Illumina, Inc. San Diego, CA, USA). Libraries were sequenced on an Illumina NextSeq using single-end 1 ⨯ 150 reads with a NextSeq 500/550 High Output v2 kit (Illumina).

Sequence Quality Control: DNA sequences were filtered for low quality (Q-Score < 20) and length (< 50), and adapter sequences were trimmed using Cutadapt. Fastq files were converted into a single fasta using shi7. Sequences were trimmed to a maximum length of 100 bp prior to alignment.

OTU (Operational Taxonomic Unit) Picking: DNA sequences were aligned to a curated database containing all representative genomes in RefSeq (NCBI Reference Sequence Database) for bacteria with additional manually curated strains. Alignments were made at 97% identity against all reference genomes. Every input sequence was compared to every reference sequence in CoreBiome’s Venti database using fully gapped alignment with BURST optimal aligner for mapping large NGS (New Generation Sequencing) data to large genome databases. Ties were broken by minimizing the overall number of unique Operational Taxonomic Units (OTUs). For the taxonomy assignment, each input sequence was assigned the lowest common ancestor that was consistent across at least 80% of all reference sequences tied for the best hit. The number of counts for each OTU was normalized to the average genome length. OTUs accounting for less than one-millionth of all species-level markers and those with less than 0.01% of their unique genome regions covered (and < 1% of the whole genome) were discarded. Samples with fewer than 10,000 sequences were also discarded. Count data were then converted to relative abundance for each sample. The normalized and filtered tables were used for all downstream analyses.

Functional Genome Content: Kyoto Encyclopedia of Genes and Genomes Orthology groups (KEGG) were observed directly using alignment at 97% identity against a gene database derived from the strain database used above. The KO table and downstream tables contain the directly observed KO counts converted to relative abundance within a sample. KOs were collapsed to level 2 and -3 KEGG pathways and KEGG Modules. In our study KEGG Level 2 (Membrane Transport, Carbohydrate Metabolism, Amino Acid Metabolism, Replication and Repair, Translation, Energy Metabolism, Metabolism of Cofactors and Vitamins, Nucleotide Metabolism, Lipid Metabolism, Transcription), KEGG Level 3 (Transporters, ABC Transporters, DNA Repair, and Recombination Proteins, Ribosome, Purin Metabolism, Peptidases, Pyrimidine Metabolism, Transcription Factors, Chromosome, Two-component System) and KEGG Enzymes (in our study n=2498) where analyzed.

Alpha and Beta Diversity: Bray-Curtis beta diversity metrics were calculated from the filtered species table and the KEGG Module and enzyme relative abundance tables using QIIME (an open-source bioinformatics pipeline for performing microbiome analysis). The Chao1 index, Shannon Index and observed OTU count (taxonomic group) were calculated using a rarefied OTU table set to the minimum depth allowed for a sample (10’000) using QIIME 1.9.1.

SMS (Shotgun Metagenomic Sequencing) has the ability to sequence the complete collection of microbial genomes present in a microbiome sample, theoretically being able to discriminate between strains that differ by a single nucleotide.

To determine the enterotype we used the enterotype classifier: http://enterotypes.org/classify.cgi, http://enterotype.embl.de/enterotypes.html (Costea et al., 2017).

### Statistics

CoreBiome supplied the data for Raw Taxonomy Tables, Filtered Taxonomy Tables, Strain Coverage Tables, Function Tables including KEGG Orthology Groups, KEGG Modules, KEGG Level 2 Pathways, KEGG Level 3 Pathways, KEGG Enzymes, Beta Diversity including Species Bray-Curtis Distance Matrix, KEGG Module Distance Matrix, KEGG Enzyme Bray-Curtis Distance Matrix, and Alpha Diversity. The statistics for downstream analysis were calculated with Analyse-it® for Excel 5.40 build 7149.20136 (Leeds, United Kingdom). For hypothesis tests Wilcoxon signed-rank test (Hodges-Lehmann Location Shift, posthoc pairwise comparisons with Tukey’s Method (False Discovery Rate correction), significance level 5%) was used, for correlations Pearson’s r.

## Results

Since the personal and healthy core native microbiome can differ between individuals due to the enterotypes (Rinninella et al., 2019),(Costea et al., 2017) we analyzed this entity in our cohort. In 1511 reference genomes Arumugam found three robust clusters (enterotypes) that are not nation or continent-specific (Costea et al., 2017). This indicates the existence of a limited number of well-balanced host-microbial symbiotic states that might respond differently to diet and drug intake. Gorvitovskaia states that the enterotypes of Prevotella and Bacteroides should be interpreted as ‘biomarkers’ of diet, lifestyle and disease state (Gorvitovskaia et al., 2016). Twenty-one of 26 of our probands belong to the enterotype 3 (Firmicutes enriched), 2 to the enterotype 1 (Bacteroides enriched), (EUC Bacteroides eggerthii 26.5%, CUC Bacteroides stercoris 39.8%) and 3 to enterotypes 2 (Prevotella enriched), (VMA 42.9%, LOH 18.22%, AAC 14.9%).

Due to the fact that sequencing can only resolve the taxonomic composition of the gut microbiome and doesn’t provide evidence of the biological functions (Ji & Nielsen, 2015) we first concentrated on the positive or negative deviations of the microbiome after three weeks of dietary supplement consumption. Figure 1 displays a stacked histogram of the bacteria found in the probands at day 0 and day 21.

**Fig. 1.**
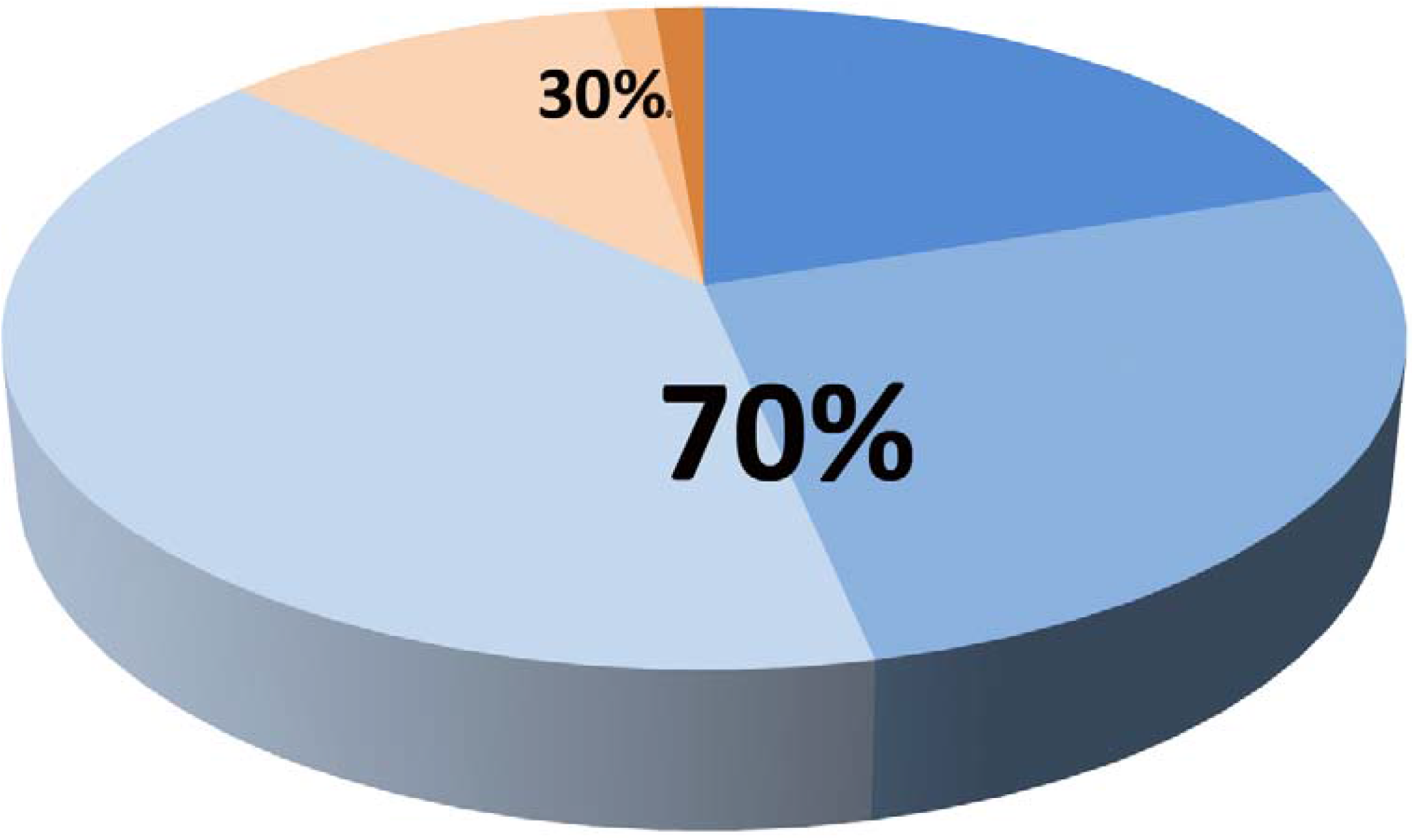
Each test person (Code at the bottom of the graph) is represented with the bacterial composition at day 0 and day 21 of the study. The variation due to the dietary supplement is apparent.

Our working hypothesis was that the dietary supplement based on baker’s yeast changes the microbiome by more than 20% within 3 weeks. Fig. 2 shows the positive and negative percentual variations in excess of 20% of the 68 bacterial species.

**Fig. 2.**
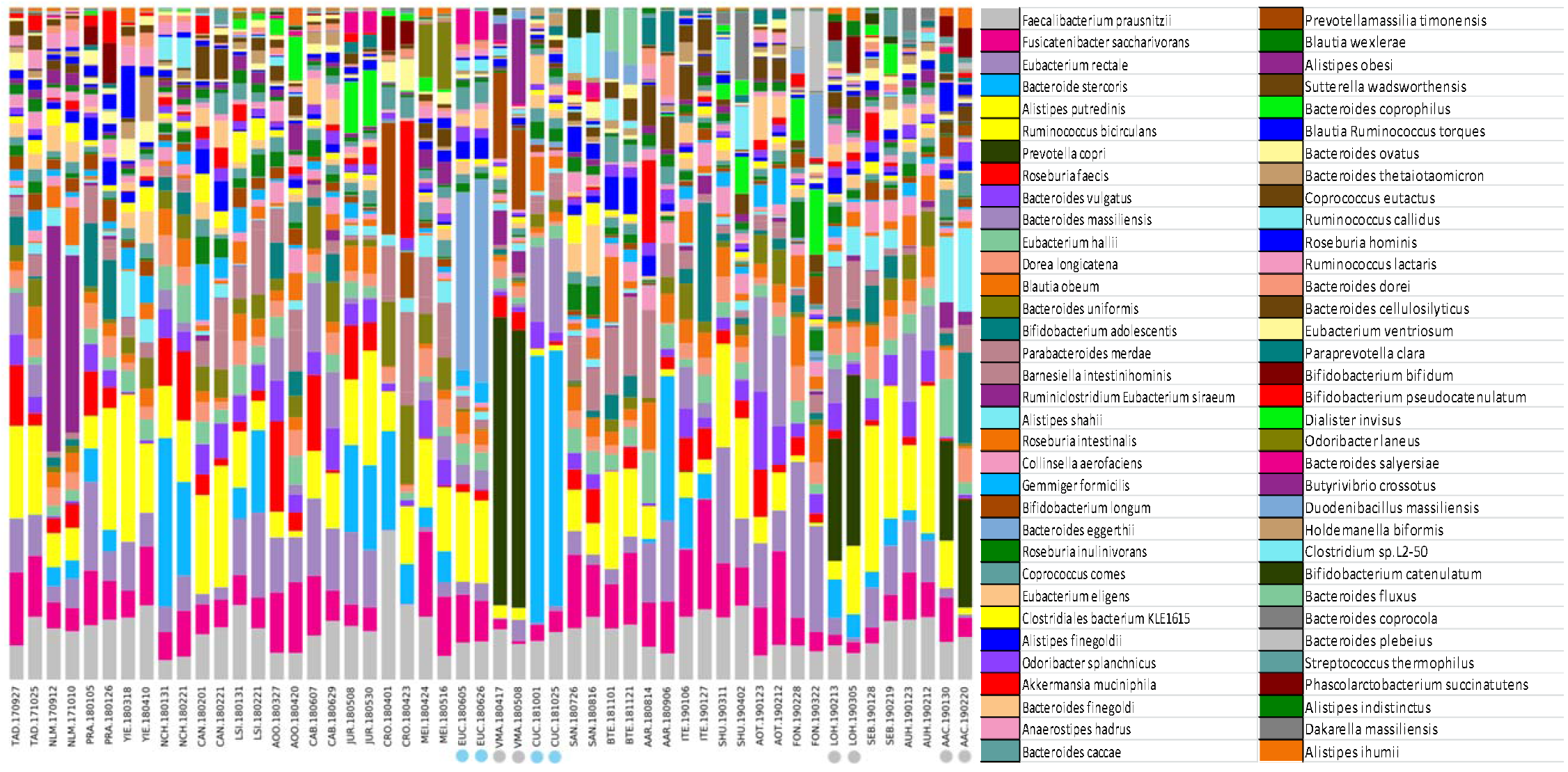
A) Thirtyone out of 68 species detected increased in excess of 20% during 3 weeks of supplement use. B) Twentythree of 68 bacteria found decreased more than 20% during supplement ingestion.

### Number of bacterial species out of 68 in the investigated cohort with an increase and decrease above 20%

From the sequence data, it is difficult to assign a specific function to a bacterium. In order to reduce the 68 bacterial species to a few known beneficial ones we selected five of the probiotic bacteria, Geirnaert described as good butyrate producers (Geirnaert et al., 2017): Anaerostipes hadrus, Faecalibacterium prausnitzii, Eubacterium hallii, Roseburia hominis and Roseburia inulinivorans. In our 26 participants, 18 showed an increase in more than three species (Fig. 3).

**Fig. 3.**
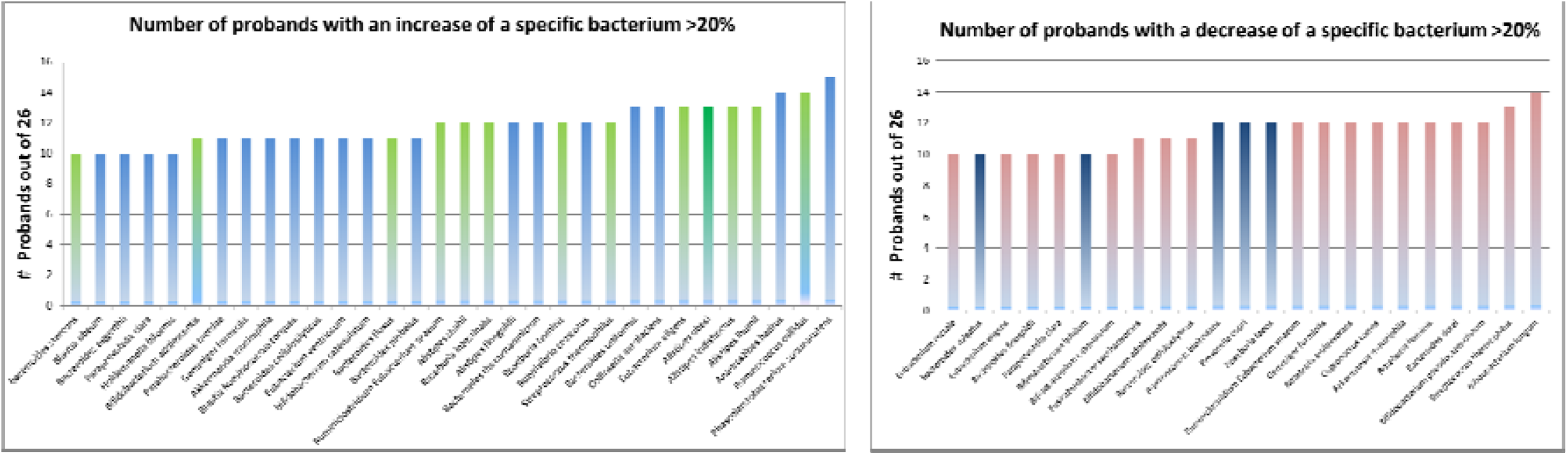
Changes in the composition of the five probiotic butyrate-producing bacteria in the studied population (n=26). Seventy percent of our cohort demonstrated a positive modification of the majority of these specific five beneficial, probiotic bacteria.

An interesting component in the evaluation of the overweight population is the Firmicutes to Bacteroidetes ratio. In the literature, a normal value is indicated as 1.6 or below (Koliada et al., 2017). In persons with a lower BMI, it can be as low as 0.7 (CI 0.6-0.7). Therefore we looked at participants with an initial F/B ratio of over 1.6. We found 12 persons in our study with an elevated ratio at the beginning. After 3 weeks the F/B ratio was lower in 14 out of 26 probands. The mean F/B ration was 1.65 before and 1.50 after treatment. This is not statistically significant but Tab.2 shows the 5 out of 12 persons starting with an increased ratio and shifting to normal values.

**Tab. 1.** Allocation of the variations in bacterial species to the individual probands (n=26)

| increase |  | decrease |  |
| --- | --- | --- | --- |
| TAD | 27 | TAD | 19 |
| NLM | 19 | NLM | 24 |
| PRA | 25 | PRA | 21 |
| YIE | 27 | YIE | 14 |
| NCH | 24 | NCH | 16 |
| CAN | 26 | CAN | 24 |
| LSI | 26 | LSI | 29 |
| AOO | 32 | AOO | 17 |
| CAB | 28 | CAB | 19 |
| JUR | 21 | JUR | 18 |
| CRO | 20 | CRO | 21 |
| MEI | 34 | MEI | 16 |
| EUC | 22 | EUC | 20 |
| VMA | 19 | VMA | 27 |
| CUC | 27 | CUC | 20 |
| SAN | 25 | SAN | 18 |
| BTE | 22 | BTE | 14 |
| AAR | 25 | AAR | 32 |
| ITE | 26 | ITE | 31 |
| SHU | 29 | SHU | 21 |
| AOT | 28 | AOT | 21 |
| FON | 27 | FON | 29 |
| LOH | 19 | LOH | 32 |
| SEB | 28 | SEB | 22 |
| AUH | 13 | AUH | 31 |
| AAC | 25 | AAC | 29 |

**Tab.2.**
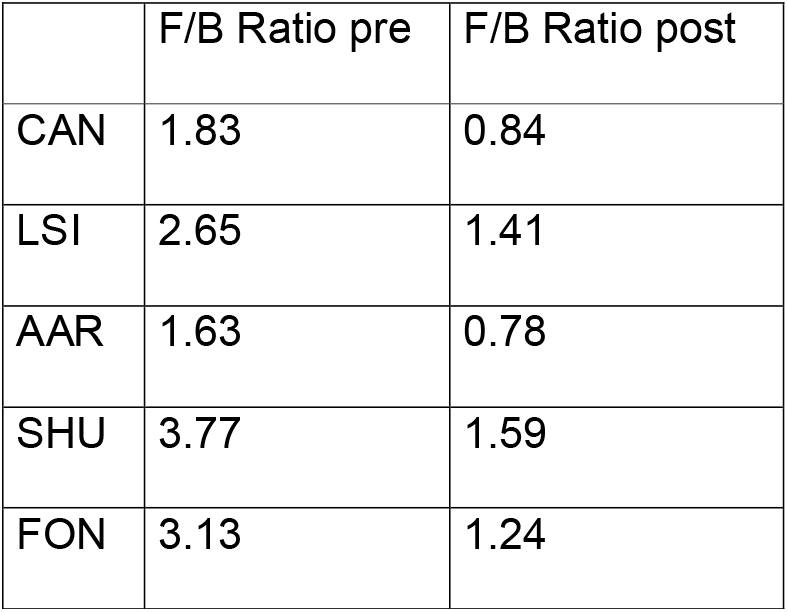
Out of 26 probands, twelve had an F/B ratio above 1.6. Five women out of these 12 reached a normal F/B ratio (below 1.6) after three weeks of taking the supplement.

### Firmicutes to Bacteroidetes Ratio

For the parameters Beta Diversity and Alpha Diversity, we didn’t find a statistically significant difference in the cohort within the 3 weeks.

## Discussion

For the understanding of the human microbiome new methods such as shotgun metagenome sequencing have brought an essential improvement. It is now possible to determine the microbiome down to the species level and to identify metabolic pathways like in KEGG.

The remaining challenge is the understanding of the cooperation of the entirety of the microbiome system. It is impossible to find direct evidence from sequencing data of the biological functions associated with the gut microbial community and there is little information about the mechanism of metabolic interactions (Ji & Nielsen, 2015). Cory Klemashevich states that to generate a beneficial effect there must be a cooperative interaction with other strains in the gut (Klemashevich et al., 2014). This makes statements about health amelioration solely from the sequence data difficult. Mircea Podar is convinced that only the cultivation of the bacteria can yield insights into their function. The human gut has as many as 1000 microbial species and the sequences cannot explain whether they contribute to digestion, immune responses, metabolism of drugs, or other processes. Moreover, also environmental exposures, microbial ecology, and human genotype confound straightforward conclusions (Turnbaugh et al., 2009). Currently, there is no consensus model of gut microbiota metabolism (Sridharan et al., 2014). It will be necessary to connect parts lists to networks in a spatial and temporal context (Raes & Bork, 2008). In the sequencing results, there is a lack of functional annotation. A large fraction of gut microbial genes is uncharacterized to date (T.S.B. et al., 2018). Amplicon sequencing typically only resolves the taxonomic composition of the gut microbiome. It is impossible to provide direct evidence of the biological functions associated with the gut microbial community(Ji & Nielsen, 2015).

Even in the homogenous group we worked with, the taxonomic composition of the gut microbiome varies greatly between individuals, due to both microbiome-intrinsic and microbiome-extrinsic factors (T.S.B. et al., 2018). Therefore the complexity prohibits a straightforward interpretation of our results apart from the genome associated entities. To make some statements concerning the changes during the supplement phase we selected 5 bacteria, which were used as probiotics in Crohn’s Disease (CD) patients (Geirnaert et al., 2017). These butyrate-producing bacteria have beneficial effects on epithelial barrier function and overall gut health. Butyrate is at the same time important to maintain gastrointestinal health because it serves as the main energy source for colonocytes, enhances epithelial barrier integrity and inhibits inflammation (Hamer et al., 2008). Seventy percent of the study participants showed an increase in the majority of these probiotic bacteria.

The enterotypes today are controversial. Nevertheless, it is interesting that 21 out of 26 probands in the studied population belong to the Firmicutes enriched group. This has an influence on the F/B ratio and therefore on the BMI.

The dietary supplement containing plasmolyzed herbal yeast fulfilled our working hypothesis of shifting the gut microbiome by more than 20 percent within 3 weeks. This shift could be linked to the yeast’s higher content of glucan and mannan. These two substances have indeed been identified to affect bacteria in the gut. Krogfelt found that mannans inserted in the gut wall facilitate the adhesion of certain bacteria. They help microbes to stay longer in the human intestinal tract by binding to human mucus or mannose sugars present on intestinal surface structures (Krogfelt, 1991). Despite the microbiome stability described for people in the age group studied, it was possible to achieve an alteration within only a three-week course of a supplement based on baker’s yeast.

## Data Availability

All data are available from the corresponding author

## Authors contributions

P.J and S.M designed the study. S.M did all the clinical work. S.C analyzed the data and contributed to the interpretation of the results. P.J analyzed the data and did the statistics. P.J was leading the writing of the manuscript. All authors provided critical feedback and helped shape the research, analysis and manuscript. All authors read and approved the final manuscript.

## Abbreviations

BMI: Body Mass Index
CD: Crohn’s Disease
F/B: Firmicutes to Bacteroidetes ratio
GALT: Gut-Associated Lymphoid Tissue
HMP: Human Microbiome Project
KEGG: Kyoto Encyclopedia of Genes and Genomes Orthology Groups
OTU: Operational Taxonomic Unit, SCFA Short-Chain Fatty Acids
SMS: Shotgun Metagenomic Sequencing

